# Community gender systems and daughter’s risk of female genital mutilation/cutting: New theory and application to Egypt

**DOI:** 10.1101/19005520

**Authors:** Kathryn M. Yount, Yuk Fai Cheong, Laurie James-Hawkins, Rose Grace Grose, Sarah R. Hayford

**Affiliations:** Hubert Department of Global Health and Department of Sociology, Emory University, Atlanta, Georgia, United States of America; Department of Psychology, Emory University, Atlanta, Georgia, United States of America; Department of Sociology and Department of Social Psychology, University of Essex, Essex, England; Department of Community Health Education, Colorado School of Public Health, University of Northern Colorado, Greeley, Colorado, United States of America; Department of Sociology, Ohio State University, Columbus, Ohio, United States of America

## Abstract

We proposed and tested a feminist social-ecological theory about daughters’ experience of female genital mutilation/cutting (FGMC) in Egypt, where over 90% of women ages 15–49 are cut. FGMC has potential adverse effects on demographic and health outcomes and has been defined as a human-rights violation. Contextual factors are important determinants of FGMC, but an integrated theory is lacking, and quantitative multilevel research is limited. We theorized that more favorable community-level gender systems, including gender norms opposing FGMC *and* expanded opportunities for women outside of the family, would be associated with a daughter’s lower risk of FGMC and would strengthen the negative association of a mother’s opposition to FGMC with her daughter’s risk of cutting. Using a national sample of 14,171 mother-daughter dyads from the 2014 Egypt Demographic and Health Survey, we estimated multilevel discrete-time hazard models to test these relationships. Community gender norms opposing FGMC had significant direct, negative associations with the hazard that a daughter was cut, but women’s opportunities outside the family did not. Maternal opposition to FGMC was negatively associated with cutting a daughter, and these associations were stronger where community opposition to FGMC and opportunities for women were greater. Results provided good support for a gender-systems theory of the multilevel influences on FGMC. Integrated, multilevel interventions that address gender norms about FGMC and opportunities for women in the community, as well as beliefs about the practice among the mothers of at-risk daughters may be needed for sustainable declines in the practice.

## Introduction

*Female genital mutilation/cutting* (FGMC) refers to “procedures involving the partial or total removal of the external female genitalia or other injury to the female genital organs for non-medical reasons” (1). The practice—especially its more extensive forms–is associated with various health and demographic outcomes, including elevated risks of an earlier age at first sex (2), sexual trauma and dysfunction (3, 4), primary infertility (5), obstetric complications (6, 7), and marital dissatisfaction, instability, and dissolution (2, 6, 8). The United Nations recognizes FGMC as a violation of human rights (1), and Sustainable Development Goal 5.3 has established a global mandate to encourage abandonment (9). Alongside legal and programmatic efforts to end FGMC (10), its prevalence has declined in some places, remains common in at least 29 mostly African countries, and remains nearly universal in eight countries, including Egypt (11, 12).

Community context influences various outcomes in childhood and adolescence related to health (13, 14), sexual behavior (15), and early marriage (16). Research on FGMC has found that community characteristics are associated with the practice among girls (17, 18), and most of this research has focused on the normative context specific to FGMC. Yet, norms about FGMC are only one aspect of a community gender system that includes structural constraints on women’s opportunities outside of the family.

We advance research on FGMC in two ways. First, we offer an integrated explanatory theory, focusing on two elements of a *community-level gender system:* the collective practice of FGMC as a *gender norm* that upholds patriarchal family systems, and the structural *opportunities for women* outside this family system. This framing highlights the salience of gender systems as an explanatory theory of demographic (19-22) and life course (23, 24) processes. Defining and testing these elements of a gender system also addresses debates about normative versus structural drivers of demographic and behavioral change (25, 26), especially for gender-related outcomes, such as skewed sex ratios at birth, girls’ excess mortality, child marriage, and lowest-low fertility. Second, we apply rigorous multilevel methods to assess how the elements of a community-level gender system are associated with the risk that a daughter is cut, directly and by moderating the influence of a mother’s FGMC attitudes and experience.

Our study site is Egypt, where FGMC remains widespread despite legal prohibitions of the practice. The 2014 Egypt Demographic and Health Survey (EDHS) provides rich data on FGMC for all women respondents and their daughters (12, 27). This study fills conceptual and empirical gaps in debates on the community influences on FGMC in what scholars have called the classic patriarchal belt of the Middle East and North Africa (MENA) region. Our framework and findings offer insights for multifaceted community interventions to further the abandonment of FGMC, and other practices harmful to women and girls.

## Background

### Prevalence, trends, and determinants of FGMC

An estimated 200 million women and girls in 30 countries have experienced FGMC (28), although estimates for the number affected vary (29). On the African continent, FGMC is practiced in at least 29 countries, with prevalence ranging from 1% in Cameroon and Uganda to 98% in Somalia (29). Across 87 national surveys for 29 African countries, the prevalence of FGMC has declined somewhat in more than half the countries; however, declines have been more pronounced in countries with low-to-moderate prevalence than in countries with near-universal prevalence, such as Egypt (11, 12, 29, 30).

Studies of the determinants of FGMC have focused on family or household characteristics—a girl’s immediate social context. In diverse settings, the daughters of mothers with more schooling (31, 32) and living in wealthier households (11, 33) less often have been cut. Other salient social contexts include religious and ethnic identities, social networks among women, and marriage markets (17, 18, 34-36).

Few studies have examined the community-level sources of variation in FGMC. Yet, even neighboring villages can harbor different attitudes and practices related to FGMC, suggesting an influence of community context on girls’ experiences (37). In the first multilevel analyses of FGMC, the community prevalence of FGMC was directly, positively associated with a girl’s risk of cutting in Kenya (17) and modified the individual-level association of religious identity on this risk in Burkina Faso (18). FGMC tends to be less common in urban than rural areas (11); however, such differences may be reduced or reversed after accounting for other factors (17, 18, 32). The influences of gender-related community characteristics are undertheorized and understudied.

### Community gender systems and FGMC: Integrating social norms and feminist theories

We propose an integrated, feminist, social-ecological framework, in which a *community-level gender system* is our explanatory model (Fig 1). We describe how two elements of this gender system*—gender norms about FGMC* and *extra-familial opportunities for women*—may influence a daughter’s risk of FGMC, directly and by conditioning the influence of maternal beliefs about and experience with the practice.

**Fig 1.**
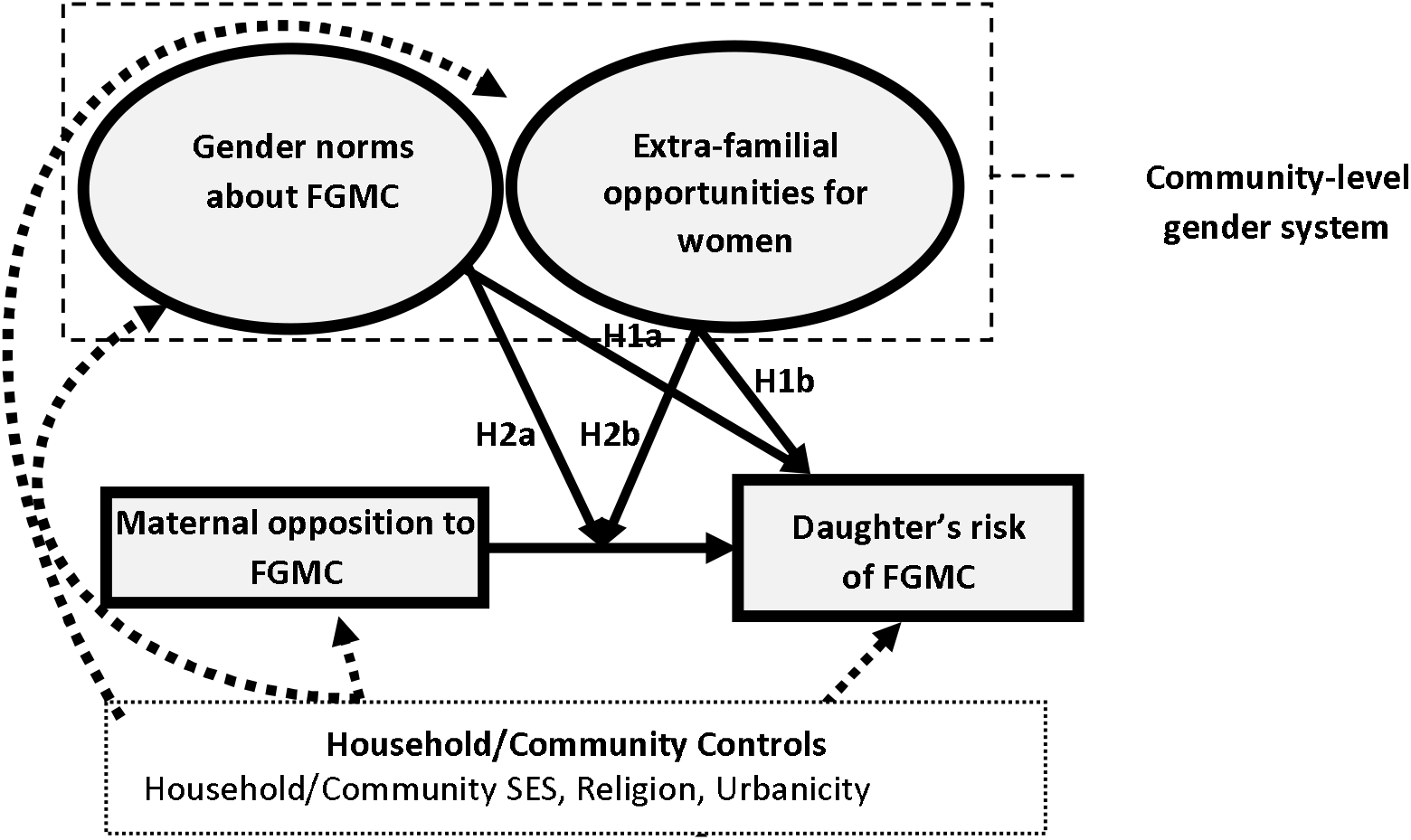
Multilevel influences on FGMC: Conceptual framework. Bolded arrows show relationships of interest. Dashed arrows show associations that we will control.

#### Community gender norms about FGMC

A *social norm* is a collective practice in a referent group that is motivated by two *social expectations* in the group: the *empirical expectation* that others practice the behavior, and the normative expectation that others think the practice should be upheld (38-41). A social norm is maintained through social influence, or expectations that compliance will be rewarded with social acceptance, and non-compliance will be punished with social rejection (41). Thus, complying with a social norm can be a marker of social belonging (42-44), and sustaining the norm is a way to uphold these social ties as distinct and valued (36).

In the case of FGMC, social norms theorists argue that the decision to cut or not to cut a daughter will depend on parental perceptions of the collective practice and of the community’s social beliefs (42, 43). A mother (usually the parent most directly involved in decisions about FGMC) may personally believe that FGMC is unnecessary or even harmful, but she may have her daughter cut if she perceives that: FGMC is practiced in her community, others think it should be practiced, and non-compliance will have negative social consequences for her daughter. That said, this mother may not have her daughter cut if she believes that her uncut daughter will be socially accepted. Social norms approaches, therefore, imply an interaction between personal beliefs and community characteristics. In communities where normative expectations are strongly enforced, opposing personal beliefs will have little influence on behavior. As normative expectations weaken, a “tipping point” will be reached where the influence of personal opposition to FGMC can be observed (42).

Social norms approaches to FGMC have some empirical support. The community-level prevalence of FGMC, a proxy for empirical expectations about the practice, has been positively associated with the risk of cutting a daughter in parts of Africa (17, 18). In communities in Egypt and parts of Africa, public denunciations of FGMC have signaled changes in normative expectations about the practice and have been thought to spur declines in prevalence (11, 45, 46). Such evidence suggests that community-level empirical and normative expectations about FGMC matter; however, the joint influences of these expectations are understudied, including whether a tipping point in community opposition to FGMC is needed for maternal opposition to have an influence on a daughter’s risk of cutting (47, 48).

A limitation of social norms theory is its cursory discussion of FGMC as integral to norms of womanhood. In many practicing societies, FGMC is performed to achieve normative expectations about sex differentiation, feminine beauty, and women’s sexuality (3, 8, 49, 50). First, women achieve social recognition by becoming less like men physically (49); thus, by removing the “male parts” and sometimes enclosing the “female parts,” FGMC “feminizes” a girl’s body (49-51). Second, FGMC helps to realize the feminine aesthetics of purity, smoothness, and cleanliness (49, 51). Third, by de-emphasizing (or controlling) women’s sexuality, FGMC accentuates women’s reproductive role (49). In these ways, the collective practice of FGMC acts as a *gender norm,* involving empirical expectations that most women are cut, normative expectations that *good women* are cut, and enforcement by requiring it for marriage in a patriarchal family system.

#### Community extra-familial opportunities for women

The gender norm of FGMC also may persist alongside *institutionalized gender inequalities* that limit women’s opportunities outside the patriarchal family (31, 52-54). These structural inequalities are multifaceted, arising in social, economic, and political domains (55). Women’s social inequality involves a lack of access to education and the means to act on their sexual and reproductive choices. Women’s economic inequality involves a lack of access to activities and institutions involving the production, distribution, and consumption of goods and services (56). Together, these institutionalized gender inequalities limit women’s access to social and economic resources and identities outside of the family, cementing women’s dependence on the security provided by (male) family members and conformity to practices that uphold this system. The strength of institutionalized inequality can be measured by the availability of *extra-familial opportunities* for women, that is, by women’s access to social and economic resources outside of the family. In lower- and middle-income settings, evidence of extra-familial opportunities for women may include higher average schooling attainments, later average ages at first marriage, lower rates of arranged marriage, and higher rates of participation in market work.

The availability of opportunities like these at the community level may allow mothers to see real alternatives to having their daughters cut (31, 39, 57). Thus, a direct association may exist between community extra-familial opportunities and a daughter’s risk of FGMC. Social norms theorists have questioned the influence of women’s extra-familial opportunities on the risk of FGMC, citing patriarchal settings where FGMC is not practiced and women’s involvement in its continuation (58). These arguments ignore feminist theory (59). First, different forms of patriarchy exist and are upheld by different gender norms, which may or may not include FGMC. Second, women’s complicity in upholding harmful practices may reflect a survival strategy under patriarchy (59), where mothers pursue the best available “options” for their daughters in the absence of real alternatives. To the extent that greater extra-familial opportunities at the community level signal a broader system of alternatives for women, they may reduce the perceived costs of violating norms favoring FGMC. Thus, an interaction also may exist between personal beliefs and community context, whereby mothers feel able not to have their daughters cut in contexts where women have sources of social and economic security outside of marriage and the marital family.

### The Egyptian context

In Egypt, FGMC remains nearly universal and a majority of women favor continuation. In 1995, 97% of ever-married women 15–49 years were cut, and the median age at cutting was 10 (60-63). Since the criminalization of FGMC in 2008, the adjusted prevalence of FGMC in girls 15–19 years declined little, from 94% in 2008 to 88% in 2014 (64). More than half of ever-married women 15–49 years supported continuation in 2008 (62%) and 2014 (58%) (64).

Here, FGMC is closely tied to expectations about gender identification, feminine beauty and purity, and control of women’s sexuality (32, 65-68). In Egyptian social systems, like many African social systems, family and community are core institutions whose interests override those of an individual. Patriarchal control and family honor are constructed through marriage and are sustained through the social and sexual control of women. By ensuring a girl’s virginity and conformity to expectations of feminine modesty and aesthetics, FGMC is thought to enhance a daughter’s prospects for a suitable marriage (32). This way, FGMC operates as a gender norm that reflects and reinforces a patriarchal family system.

In recent decades, minority Christian groups have begun to abandon FGMC while Islamist groups have promoted FGMC as part of authentic Muslim womanhood (36). Thus, higher percentages of Muslim than Christian women practice FGMC, intend to cut their daughters, and view FGMC as desired by religion, “good” for the girl, cleansing or purifying, and protective against sexual transgressions (32). Findings from the 2014 Survey of Young People in Egypt confirm that Christian women and men are much less likely than Muslim women and men to report favorable views about the practice (69). Thus, the emergence of FGMC as a symbol of authentic Muslim womanhood has transformed the practice from a widespread gender norm to a gender norm aligned more narrowly with Muslim identity (36).

In this context, opportunities for women outside the family remain limited. In 1981, Egypt ratified the Convention on the Elimination of All Forms of Discrimination against Women (CEDAW), but with reservations to the article on equality in marriage and family life (70). Child and consanguineous or blood marriage remain common, and divorce remains rare (27). Namely, more than 17% of women 20–24 years were married as children, and among the poorest and least educated women, the median age at first marriage remains less than 19 years (27). One third (31%) of ever-married women report consanguineous marriage, of which more than half are first-cousin marriages (27). Child marriage and cousin marriage have been associated with women’s lower influence in household decisions (57, 71), which in turn, has been related to a woman’s higher intent to cut a daughter (72). Although women and men legally can seek divorce under certain conditions (73), divorce remains stigmatized and rare, with less than 3% of women 35–49 years divorced in 2014 (27). Alongside women’s social dependence on marriage, their economic dependence also remains high, as evidenced by lower rates than men of high school completion (33% versus 38%) (27) and market work, especially after marriage and childbearing (74, 75).

### Summary and hypotheses

In sum, an integrated feminist social-ecological theory situates the decision to cut or not to cut a daughter within a *community gender system*. This system includes *gender norms* that uphold or challenge patriarchal marriage systems and *extra-familial opportunities* available or not available to women. Both elements of this gender system will directly influence a daughter’s risk of FGMC and will condition the influence of a mother’s opposition to FGMC on the risk that her daughter is cut. Specifically:

*H1*. A daughter will have a lower adjusted hazard of FGMC in communities *(a)* that have a lower prevalence of FGMC and more strongly oppose the practice, and *(b)* where women have more extra-familial opportunities.

*H2*. A mother’s opposition to FGMC and non-cut status will be associated with a lower hazard of her daughter being cut in communities *(a)* that have a lower prevalence of FGMC and more strongly oppose the practice, and *(b)* where women have more extra-familial opportunities. In communities with less favorable gender systems, maternal opposition to FGMC will not be associated with a daughter’s risk of FGMC.

## Method

### Sample and data

The data source for this analysis was the 2014 EDHS. The DHS collects nationally representative data from women of reproductive age and their households on household socioeconomic conditions, women’s marriage and birth histories, schooling and work, and FGMC, among other data. The DHS typically use a two-or three-stage probability sampling design, in which small geographic areas are selected, followed by households and respondents within households. These small-sample areas can serve as effective proxies for villages or neighborhoods (76).

The Ministry of Health and Population provided ethical approval for the 2014 EDHS, and respondents provided verbal informed consent to participate (77). The data for this analysis were publically available, de-identified secondary data provided by permission from Measure DHS. Emory University’s Institutional Review Board, therefore, considered this analysis exempt.

The 2014 EDHS included 883 sample clusters after excluding 42 (<1%) in Sinai for security reasons. On average, 25 households per cluster were sampled. Within each household, all ever-married women 15–49 years were interviewed, for a total sample size of 21,762 women. Because our outcome was the FGMC experience of daughters, we treated the mother-daughter dyad as the individual-level, or level-one, unit of analysis. The sample cluster was the community-level, or level-two, unit of analysis. For mothers with more than one daughter, we randomly selected one daughter for the analysis. Women with no daughters were included only in constructing level-two measures (see below). After dropping mother-daughter dyads with missing data (n=26), the final sample size was 14,171 mother-daughter dyads within households nested in 881 communities. On average, these clusters included 16 mother-daughter dyads (ranging from 1 to 78 across clusters, with a standard deviation of 8). Less than 1% of clusters included only 1 woman, 7.5% had ≤5 women, and 27% had ≤10 women. For primary exposure variables, the community-level sample averaged 24 ever-married women 20–49 years per cluster (ranging from 1 to 127 across clusters, with a standard deviation of 13).

The 2014 EDHS collected three general types of data on FGMC. All ever-married women 15–49 years self-reported their experience with FGMC, including type and timing, and their personal attitudes about FGMC. All ever-married mothers 15–49 years reported their daughters’ experiences with FGMC, including age of the event for cut daughters.

### Main variables

#### Outcome: Daughter’s FGMC status

Our outcome was the *daughter’s experience* of FGMC. For each daughter aged 20 years or younger, the mother reported whether her daughter was cut. If the response was “yes,” the mother reported the daughter’s age at cutting in years.

#### Individual (level-one) exposure

A measure for *maternal opposition to FGMC* was derived from a factor analysis of four items that captured non-cutting and disfavorable attitudes about FGMC. The first item captured whether the mother reported to be *uncut* (0=no, 1=yes). Inclusion of cutting status was justified conceptually and empirically. First, the cut or uncut status of the mother arguably captured a preference for the practice among her own parents (78, 79). Second, evidence from Egypt has shown a strong, positive association of maternal FGMC status with maternal intent to cut a daughter and a daughter’s risk of being cut (32). We treated this item as dichotomous because self-reports of any versus no cutting are reasonably reliable (60, 80, 81). The second and third items captured whether the mother believed FGMC was *not* required by religion (0=no, 1=yes) and should *not* continue (0=no, 1=yes). For both questions, the original response option ‘don’t know’ was recoded as 0 so that unfavorable attitudes about FGMC were clearly distinguished. Because of the strong association of Christian religion with unfavorable attitudes about FGMC and non-cutting in Egypt (32, 36), we added a fourth item for whether the mother was Christian (=1) or Muslim (=0). In random split-half samples, we used exploratory factor analysis (EFA) and confirmatory factor analysis (CFA) to assess whether these items reflected a unidimensional construct. A single-factor CFA model with all four items had an adequate fit to the data (factor loadings 0.53–0.98, Root Mean Squared Error of Approximation [RMSEA]=0.02, Comparative Fit Index [CFI]=0.99, Tucker-Lewis Index [TLI]=0.99, α = 0.65) (82-84).

#### Community (level-two) exposures

We considered four items to capture *community-level gender norms opposing* FGMC. Items were created using data from all ever-married women 20–49 years in the community, a separate, non-overlapping, cohort from the daughters ages 0–20 on which the outcome was observed. One measure of empirical expectations about FGMC was the proportion of women in the community who were *uncut*. Two measures of normative expectations about FGMC captured the proportions of women agreeing that religion does *not* require FGMC and that it should *not* continue. The proportion of women who were Christian was considered for the reasons described, above. In random split-half samples of communities, we performed EFA and CFA to assess whether these four items reflected a unidimensional construct. The proportion Christian did not adequately load and was dropped. A single-factor CFA model with the remaining three items (85) had an adequate fit to the data (factor loadings 0.72–0.99, RMSEA=0.12, CFI=0.93, TLI= 0.90, α = 0.89). Although the 2014 EDHS had a men’s sample, we used women’s responses to capture community gender norms about FGMC because men often report less favorable attitudes about FGMC than do women (86, 87).

We also created a factor score for *women’s community-level extra-familial opportunities*. This score was based on the proportions of ever-married women ages 20–49 in the community who had completed secondary school or higher, had first lived with a spouse at age 18 or older, had married a non-cousin, and had worked for cash or kind in the prior year. We considered adding a measure for women’s ownership of land, but less than two percent of ever-married women 15–49 years owned land in 2014 (77). We also considered aggregate scores for women’s freedom of movement and influence in household decisions; however, questions on freedom of movement were not asked in the EDHS 2014, and measures for freedom of movement and decision-making capture women’s agency (54), not alternatives to marriage, a core element of our theory. In Egypt, women’s age at first marriage, schooling, and market work are positively associated with their agency (57). We performed an EFA and a CFA in random split-half samples. A single-factor CFA model with all four items had an adequate fit to the data (factor loadings 0.57–0.84, RMSEA=0.12, CFI=0.93, TLI=0.90, α=0.68).

#### Level-one control variables

We controlled for mother-daughter characteristics that have been associated with a daughter’s risk of experiencing FGMC. We included the daughter’s birth year in calendar years to control for cohort differences in FGMC risk (64), daughter’s birth order among daughters (32), mother’s age in years (18, 32), father’s schooling attainment (1=completed secondary or higher, 0=less than completed secondary) (32, 88), and a DHS-created household wealth score dichotomized to capture the wealthiest two quintiles (top 40%) versus the bottom 60% of households (29, 32).

We also controlled for a composite level-one measure for maternal opportunities. This measure was derived from a factor analysis of four items: maternal reported age at first residence with a spouse (0=17 or younger, 1=18 or older), non-cousin marriage (0=no, 1=yes), schooling attainment (0=less than completed secondary, 1=completed secondary or more), and work outside the home for cash or kind in the prior year (0=no, 1=yes). In random split-half samples, we performed EFA and CFA to assess whether these items reflected a unidimensional construct. Non-cousin marriage loaded poorly and was dropped. A single-factor CFA model with the remaining three items had an adequate fit to the data (factor loadings 0.40–0.99, RMSEA=0.02, CFI=0.99, TLI=0.99, α=0.48).

#### Level-two control variables

We controlled for three community-level characteristics that could confound the relationship between community norms, women’s opportunities, and a daughter’s risk of FGMC: the proportion of households in the wealthiest two quintiles, defined above (17); urbanicity using DHS definitions for urban versus rural locality (11, 32); and the proportion of ever-married women 15–49 years who were Muslim (11, 36). All ever-married women 15-49 years were used to construct the last two variables to control for a community’s religious and urban composition with all available data.

#### Analyses

The analysis was of de-identified secondary data, so the study was exempt from IRB review. We began with descriptive analyses showing the characteristics of mother-daughter units in the sample and variation in the characteristics of these units according to community-level opposition to FGMC. All level-one descriptive statistics were weighted, and robust standard errors were estimated to account for the stratified, cluster-sample design. We also presented point estimates and ranges for community-level characteristics (unweighted because the DHS does not provide cluster-level weights).

We used multilevel discrete-time hazard models to estimate associations of a daughter’s hazard of FGMC with factor scores for maternal FGMC attitudes and status, community FGMC norms, and community opportunities for women (89-91). Event-history models predict the likelihood of FGMC at each age, accounting for the possibility that girls uncut at the survey may be cut in the future. To facilitate model estimation, we used age-risk-sets, or multi-year periods of exposure, rather than person-years as the unit of analysis. The multilevel structure of the models accommodates the hierarchical nature of the data and avoids the problem of biased estimates of precision that result from ignoring clustering within communities. All models were estimated using STATA 14.1 (92).

Let *η*_*ijt*_ denote the log-odds of the baseline hazard of FGMC for the daughter of mother *i* living in community *j* at daughter’s age *t*. Let *A*_*ijt*_ denote the daughter’s age risk set, specified as T1=0–1 years, T2=2–11 years, T3=12–16 years, and T4=17–20 years to capture major age-specific risk sets for FGMC in this setting (89-91, 93). Since the model estimated was a limited dependent variable hazard model, we included all four age risk sets capturing ages 0–20 to provide maximum information to estimate the hazard for each age risk set. Although the median age of cutting in 2014 in Egypt was 10.5 years among ever-married women ages 15–49 (77), the probabilities of cutting at individual ages before age 10 ranged from .01 to .05 and were substantial enough that we did not truncate the sample of girls to those ages 10 to 20. Let *FGMC*_*ij*_ denote the mother’s FGMC attitudes and status, *N*_*j*_ the community FGMC norms, and *O*_*j*_ the community extra-familial opportunities for women. A general two-level discrete-time hazard model can be depicted as:

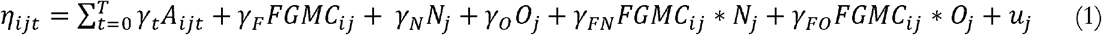

Equation (1) shows *η*_*ijt*_ as a function of the age risk set t of the daughter of mother *i* in community *j*; main exposure variables at level-one and level-two and their interactions; and random community-specific contributions, *u*_*j*_, which are assumed to be normally distributed with variance δ. The γ_*t*_’s identify the logit hazard curve, and γ_*F*_, γ_*N*_, γ_*O*_, γ_*FN*_, and γ_*FO*_, identify the effects associated with the exposures and their interactions. Controls at level-one and level-two are added to Eq. (1) but are not interpreted (94). Level-one variables were cluster-mean centered, and level-two variables were grand-mean centered. The magnitude and significance of coefficients of the community-level attributes provided tests of hypothesis 1, regarding direct community effects. The magnitude and significance of coefficients of the interaction terms provided tests of hypothesis 2, regarding conditional community effects.

To assess the “tipping point” hypothesis (42, 43, 95), significant cross-level interactions were probed to detect the range of values for the community exposures for which maternal opposition to FGMC was significantly associated with her daughter’s FGMC risk (96). Social norms theory suggests that when community *support* for FGMC is high, even mothers who oppose the practice will have their daughters cut; conversely, when community *opposition* to FGMC is high, and a new and self-sustaining norm *against* FGMC emerges, the influence of maternal opposition to FGMC is observed. The “tipping point” refers to the boundary levels of community opposition to FGMC or community opportunities for women at which maternal opposition to FGMC is related significantly to a daughter’s hazard of FGMC.

To assess the proportional odds assumption of these models, that all covariates have the same effects for all age risk sets (93), we tested the significance of two-way interactions of all covariates with the age-risk-set indicators (available on request). Our final models controlled for two covariates with non-proportional effects, maternal opposition to FGMC and daughter’s birth year, by retaining their interaction terms in the final model. For Muslim and living in an urban area, we attempted to test the proportional error assumption that the logit-hazard curves of the communities were parallel to one another, holding constant all predictors (93); however, the algorithm could not produce estimates, perhaps because less than 1% of observations on these two variables were in the last age-risk set.

We did not apply multilevel sampling weights to account for the unequal probabilities of cluster and participant selection because the DHS does not provide cluster-level sampling weights (97). In a similar single-level analysis that incorporated individual-level sampling weights and stratification information, inferences were robust to the inclusion or exclusion of weights.

## Results

### Characteristics of the mother-daughter sample

On average, daughters were eight years old at the time of the survey (born in 2005), and either the first- or second-born daughter (Table 1). At the time of the survey, 20% of daughters had been cut, and, as expected, this prevalence was monotonically higher across age groups of daughters, with less than 1% of 0–1 year-olds, 6% of 2–11 year-olds, 49% of 12–16 year-olds, and 65% of 17–20 year-olds cut.

**Table 1.**
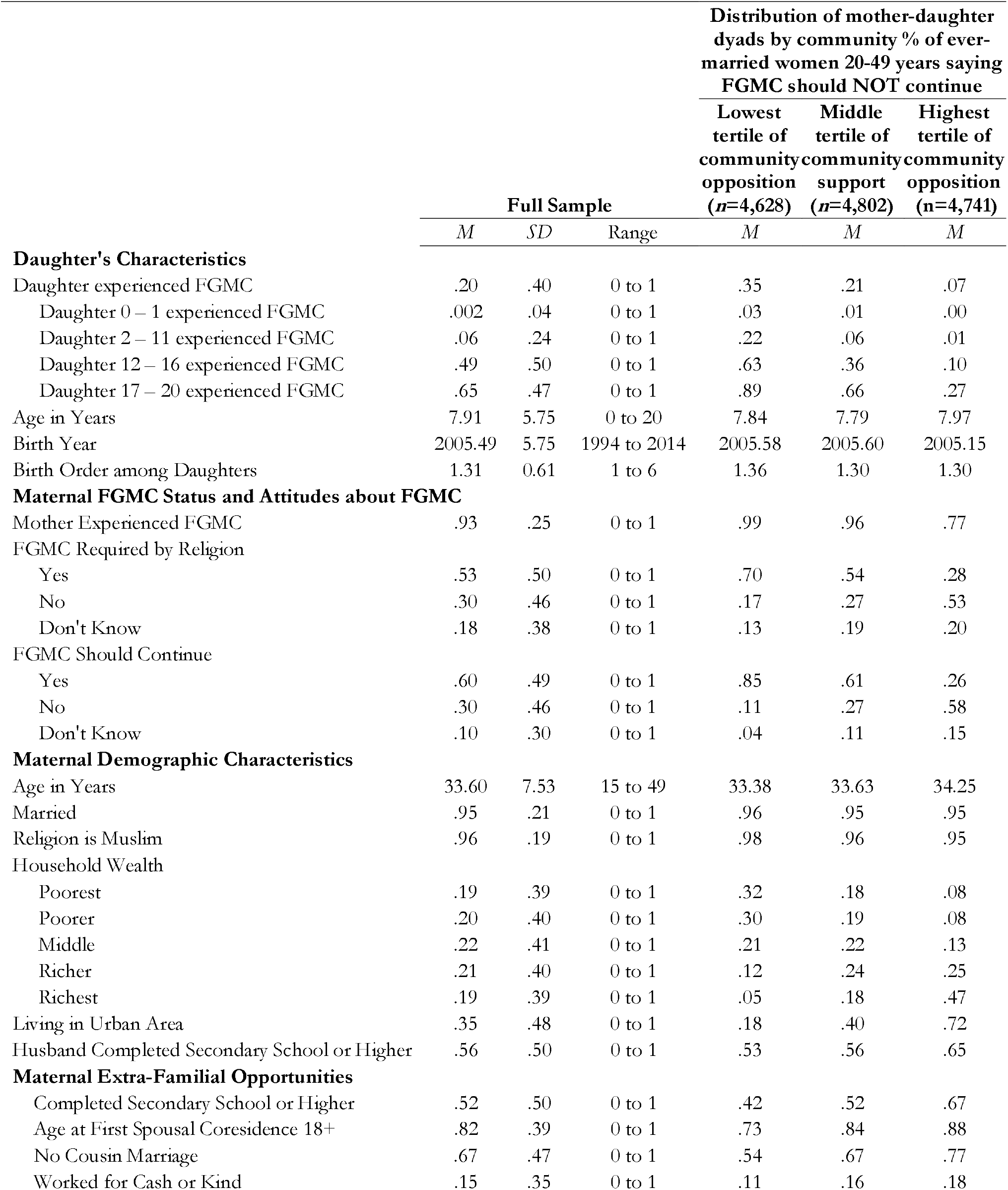
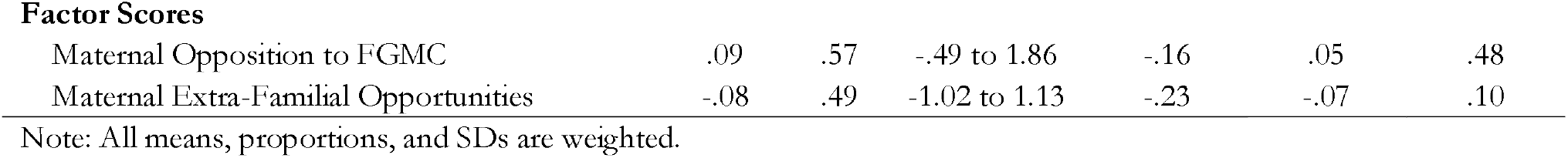
Characteristics of Ever-Married Mothers 15–49 Years and their Daughters 0–20 Years, 2014 Egypt Demographic and Health Survey (*N* = 14,171 mother-daughter dyads).

|  | Full Sample |  |  | Distribution of mother-daughter dyads by community % of ever-married women 20-49 years saying FGMC should NOT continue |  |  |
| --- | --- | --- | --- | --- | --- | --- |
|  |  |  |  | Lowest tertile of community opposition (*n*=4,628) | Middle tertile of community support (*n*=4,802) | Highest tertile of community opposition (*n*=4,741) |
|  | *M* | *SD* | Range | *M* | *M* | *M* |
| **Daughter's Characteristics** |  |  |  |  |  |  |
| Daughter experienced FGMC | .20 | .40 | 0 to 1 | .35 | .21 | .07 |
| Daughter 0 – 1 experienced FGMC | .002 | .04 | 0 to 1 | .03 | .01 | .00 |
| Daughter 2 – 11 experienced FGMC | .06 | .24 | 0 to 1 | .22 | .06 | .01 |
| Daughter 12 – 16 experienced FGMC | .49 | .50 | 0 to 1 | .63 | .36 | .10 |
| Daughter 17 – 20 experienced FGMC | .65 | .47 | 0 to 1 | .89 | .66 | .27 |
| Age in Years | 7.91 | 5.75 | 0 to 20 | 7.84 | 7.79 | 7.97 |
| Birth Year | 2005.49 | 5.75 | 1994 to 2014 | 2005.58 | 2005.60 | 2005.15 |
| Birth Order among Daughters | 1.31 | 0.61 | 1 to 6 | 1.36 | 1.30 | 1.30 |
| **Maternal FGMC Status and Attitudes about FGMC** |  |  |  |  |  |  |
| Mother Experienced FGMC | .93 | .25 | 0 to 1 | .99 | .96 | .77 |
| FGMC Required by Religion |  |  |  |  |  |  |
| Yes | .53 | .50 | 0 to 1 | .70 | .54 | .28 |
| No | .30 | .46 | 0 to 1 | .17 | .27 | .53 |
| Don't Know | .18 | .38 | 0 to 1 | .13 | .19 | .20 |
| FGMC Should Continue |  |  |  |  |  |  |
| Yes | .60 | .49 | 0 to 1 | .85 | .61 | .26 |
| No | .30 | .46 | 0 to 1 | .11 | .27 | .58 |
| Don't Know | .10 | .30 | 0 to 1 | .04 | .11 | .15 |
| **Maternal Demographic Characteristics** |  |  |  |  |  |  |
| Age in Years | 33.60 | 7.53 | 15 to 49 | 33.38 | 33.63 | 34.25 |
| Married | .95 | .21 | 0 to 1 | .96 | .95 | .95 |
| Religion is Muslim | .96 | .19 | 0 to 1 | .98 | .96 | .95 |
| Household Wealth |  |  |  |  |  |  |
| Poorest | .19 | .39 | 0 to 1 | .32 | .18 | .08 |
| Poorer | .20 | .40 | 0 to 1 | .30 | .19 | .08 |
| Middle | .22 | .41 | 0 to 1 | .21 | .22 | .13 |
| Richer | .21 | .40 | 0 to 1 | .12 | .24 | .25 |
| Richest | .19 | .39 | 0 to 1 | .05 | .18 | .47 |
| Living in Urban Area | .35 | .48 | 0 to 1 | .18 | .40 | .72 |
| Husband Completed Secondary School or Higher | .56 | .50 | 0 to 1 | .53 | .56 | .65 |
| **Maternal Extra-Familial Opportunities** |  |  |  |  |  |  |
| Completed Secondary School or Higher | .52 | .50 | 0 to 1 | .42 | .52 | .67 |
| Age at First Spousal Coresidence 18+ | .82 | .39 | 0 to 1 | .73 | .84 | .88 |
| No Cousin Marriage | .67 | .47 | 0 to 1 | .54 | .67 | .77 |
| Worked for Cash or Kind | .15 | .35 | 0 to 1 | .11 | .16 | .18 |
| Factor Scores |  |  |  |  |  |  |
| Maternal Opposition to FGMC | .09 | .57 | -.49 to 1.86 | -.16 | .05 | .48 |
| Maternal Extra-Familial Opportunities | -.08 | .49 | -1.02 to 1.13 | -.23 | -.07 | .10 |

Note: All means, proportions, and SDs are weighted.

On average, mothers were 34 years old, married (95%), Muslim (96%), living in a rural area (65%), and had a spouse who completed secondary school (56%) (Table 1). A majority of mothers had completed secondary school (52%) and were age 18 or older when they began living with a spouse (82%). Only 15% had worked for cash or kind in the prior year. All mothers had heard of FGMC, most were cut (93%), and a majority felt FGMC was required by religion (53%) and should continue (60%).

The characteristics of daughters and mothers differed across communities with the lowest, middle, and highest opposition to FGMC, measured as tertiles of the community proportion of ever-married women 20–49 years believing that FGMC should not continue (Table 1). Only 7% of daughters were cut in communities with the highest opposition to FGMC, compared to 21% and 35% of daughters being cut in communities where opposition to FGMC was moderate or low, respectively. Also, the percentages of mothers who were cut (99% to 96% to 77%), believed that FGMC was required by religion (70% to 54% to 28%), and believed that FGMC should continue (85% to 61% to 26%) were monotonically lower with higher community opposition to FGMC (Table 1). Maternal secondary schooling, work for cash or kind, and first spousal co-residence at age 18 or older were more common in communities with high opposition to FGMC than in communities with low or moderate opposition to FGMC. Mothers whose spouse had the least schooling, who lived in the poorest households, and who lived in rural areas tended to be in communities with the lowest opposition to FGMC (Table 1).

### Characteristics of communities

About half of communities (51%) were urban (Table 2). In the average community, a majority of women were Muslim (96%), first lived at age 18 or older with a spouse (85%), married a non-cousin (68%), had completed secondary school (57%), and lived in the wealthiest 40% of households (52%); however, a minority of women had worked for cash or kind in the prior year (16%), were uncut (11%), believed that FGMC should stop (36%), and believed that religion did not require FGMC (34%).

**Table 2.**
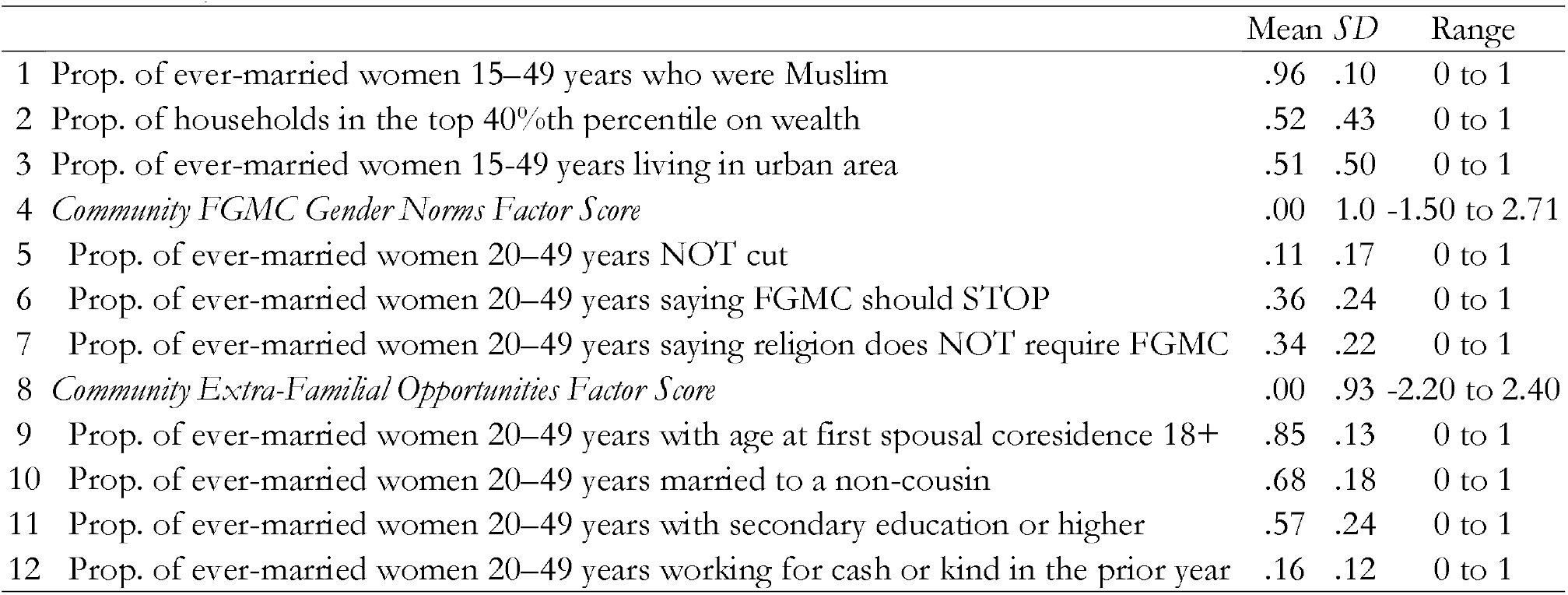
Level-Two Community Characteristics, 2014 Egypt Demographic and Health Survey (*N* = 881 Communities)

### Multilevel discrete-time hazard results

Results of multilevel discrete-time hazard models for a daughter’s risk of FGMC are in Table 3. Model 1, the unconditional model, includes dummy variables for daughter’s age risk sets. Model 2 adds level-one and level-two explanatory variables. Model 3 adds hypothesized cross-level interactions. Model 4 adds level-one and level-two control variables and significant two-way interactions between age risk sets and covariates to adjust for the non-proportional effects of the latter.

**Table 3.**
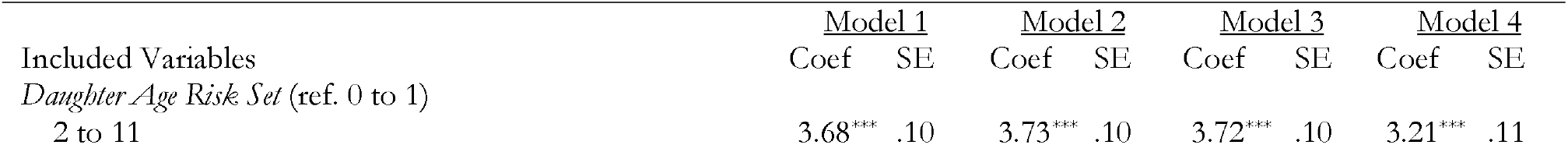
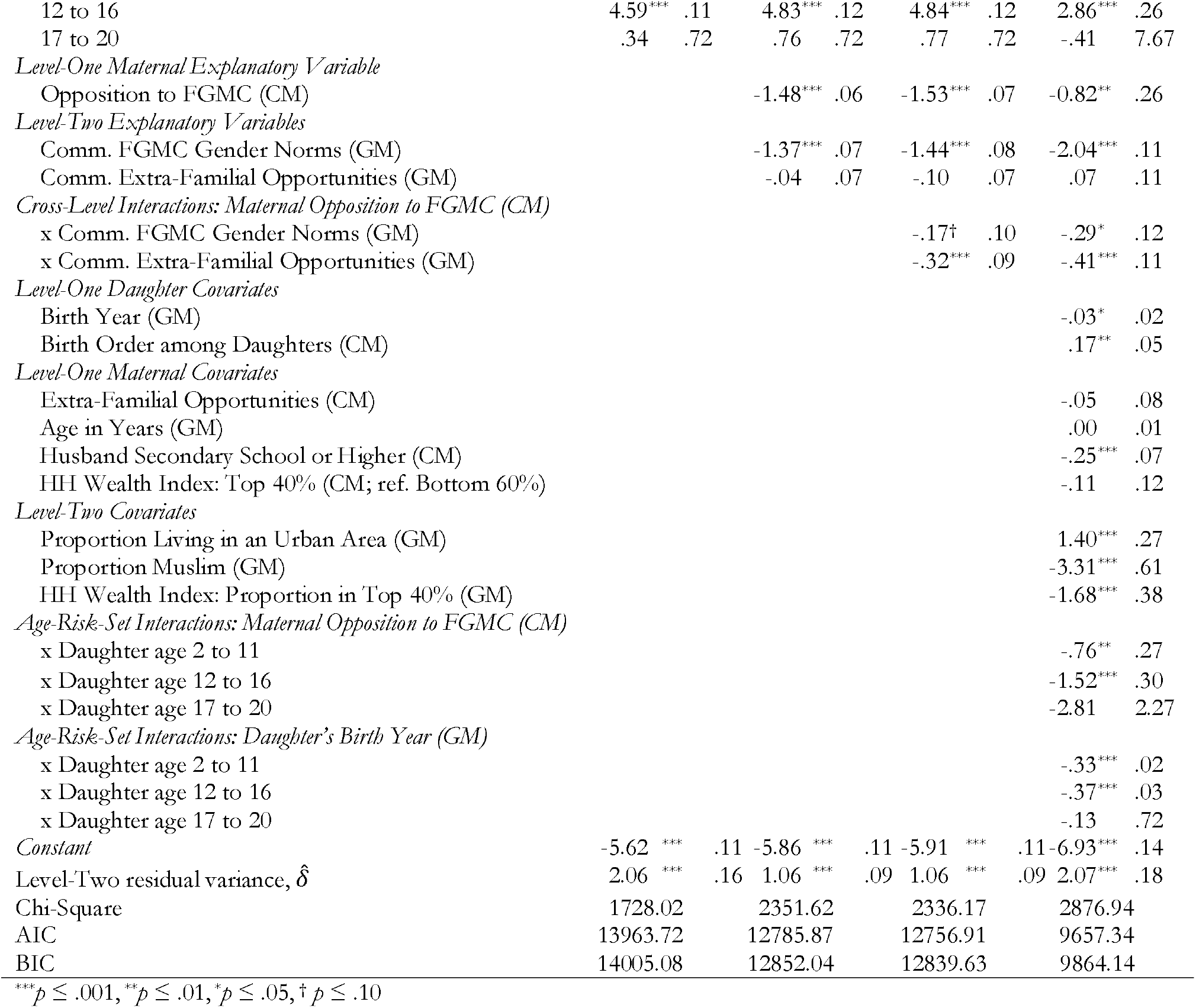
Two-Level Discrete-Time Hazard Models Predicting Daughter’s Risk of FGMC, 2014 Egypt Demographic and Health Survey (*N* = 14,171 mother-daughter pairs).

In Models 1–4, the hazard of FGMC among daughters was higher among 2–11 and 12–16-year-olds than among 0–1-year-olds (the reference group). The hazard was comparable between 17– 20 and 0–1 year-olds. In unconditional models with single-year age dummies (available on request), the hazard of FGMC appeared to peak at age 12.

In Model 2, consistent with hypothesis 1 regarding community influences, community FGMC norms were negatively associated with the hazard that a daughter was cut. Specifically, a daughter living in a community with less favorable FGMC norms among women had a lower hazard of experiencing FGMC. However, living in a community with more extra-familial opportunities for women had no significant direct association with the hazard that a daughter was cut. Stronger maternal opposition to FGMC (her non-cutting and more unfavorable attitudes) was associated with a lower hazard of FGMC for her daughter.

In Model 3, both cross-level interactions were at least marginally significant, meaning the association of maternal opposition to FGMC with her daughter’s hazard of FGMC depended on community FGMC norms and opportunities for women. In Model 4, with controls, associations of interest were robust, and sometimes larger, such as the interaction of community FGMC norms with maternal opposition. This finding suggests that some covariates, such as the community proportions urban, Muslim, and with the wealthiest 40% of households, may be negative confounders.

Based on Model 4, we graphed the simple slope (or estimated coefficient) and its 95% confidence bands for the relationship between a mother’s level of opposition to FGMC and her daughter’s predicted logit hazard of FGMC, conditional on the level of community opposition to FGMC (Fig 2). These estimates were generated for the age-risk-set 2–11, with the highest risk of cutting, and other covariates in the model were held constant at zero. Fig 2 shows that the slope for maternal opposition to FGMC was more negative in communities with stronger opposition to FGMC. Specifically, maternal opposition to FGMC was more protective against a daughter’s risk of FGMC in communities that more strongly opposed FGMC. Using the online tool of Preacher and colleagues (96), we found that the relationship of maternal opposition to daughter’s FGMC was significant in communities where opposition to FGMC was 3.18 SD below the grand mean or higher (Fig 2). Thus, in communities where opposition to FGMC was *weakest* (3.18 SD below the grand mean or lower), maternal opposition to FGMC was unrelated to her daughter’s FGMC hazard, consistent with the idea that opposition to FGMC must reach a “tipping point” before mothers who oppose FGMC enact their preferences. In this sample, only one community would have a value less than 3.18 SD below the community grand mean of FGMC opposition, however.

**Fig 2:**
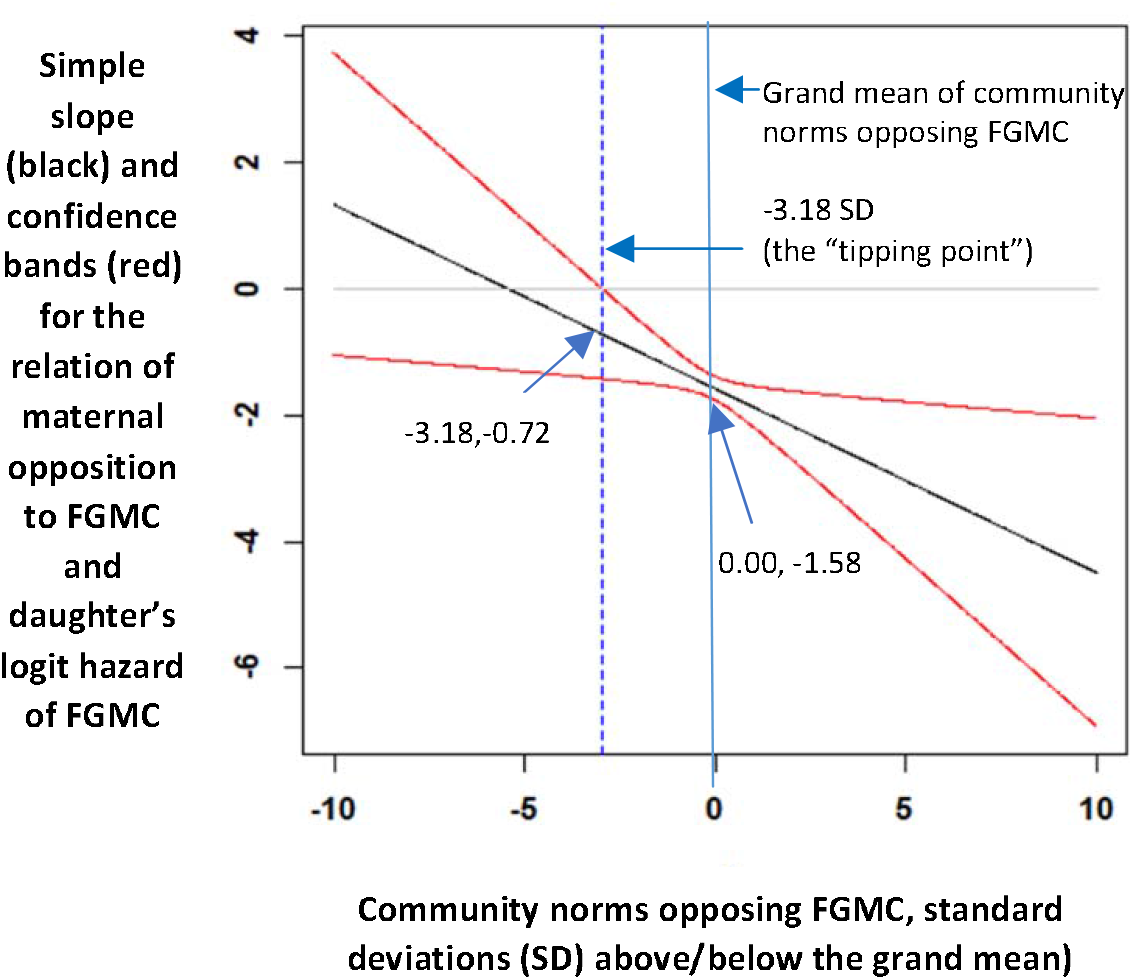
Predicted logit hazard of FGMC for the age-risk-set 2–11 years, Egypt 2014. Estimated logit hazards are generated from Table 3, Model 4 as a function of maternal opposition to FGMC at different levels of community norms opposing FGMC

Fig 3 shows a similar plot for the relationship of a mother’s level of opposition to FGMC with her daughter’s predicted hazard of FGMC across communities with different levels of opportunities for women, for the age-risk-set 2–11. The downward slope in the plot shows that the coefficient for maternal opposition to FGMC, again, was more negative in communities with more opportunities for women. Specifically, maternal opposition to FGMC was more protective against a daughter’s risk of FGMC in communities where women had more opportunities outside of marriage. The coefficients for maternal opposition to FGMC were significant in communities with extra-marital opportunities 2.79 SD below the mean level of opportunity or higher. Thus, in addition to a critical mass of opposition to FGMC, as predicted by social norms theory, there appears to be a “tipping point” of women’s opportunity, as well. In communities below this tipping point, mothers who oppose FGMC are less likely to enact their preferences. Again, in this sample, only two communities would have a value less than 2.79 SD below the community grand mean of women’s opportunity. Results for the other age risk sets are consistent.

**Fig 3:**
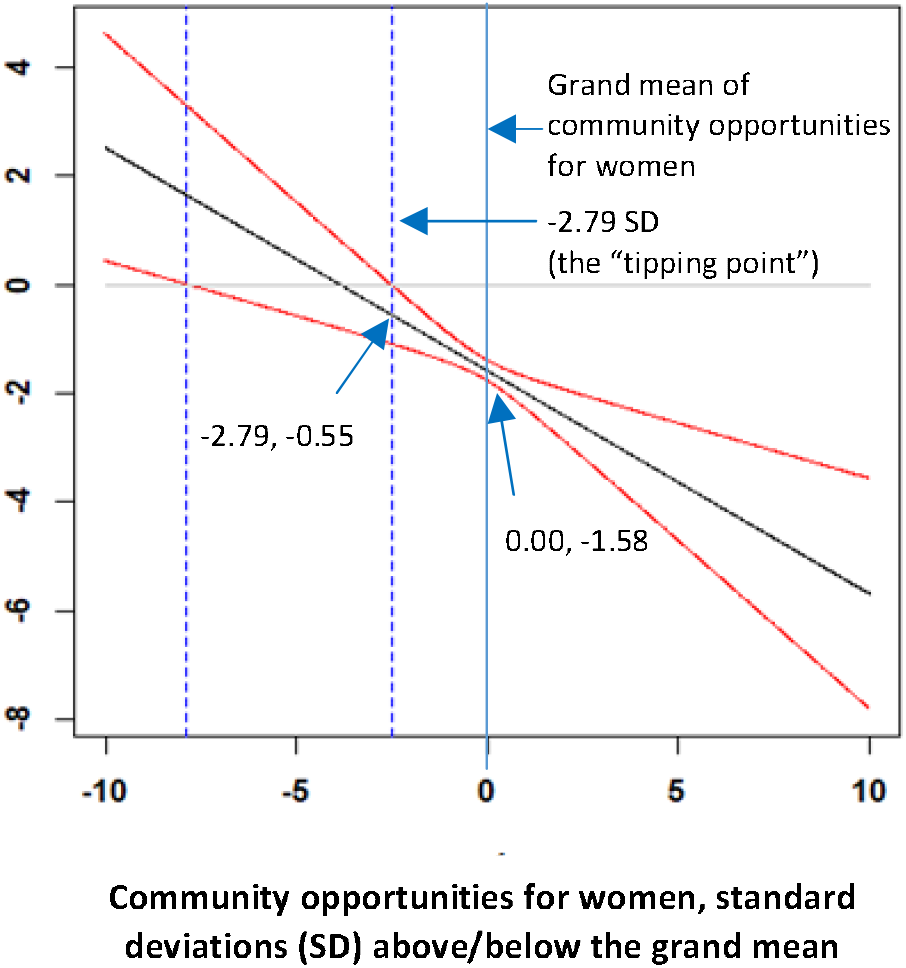
Predicted logit hazard of FGMC for the age-risk-set 2–11 years, Egypt 2014. Estimated logit hazards are generated from Table 3, Model 4 as a function of maternal opposition to FGMC at different levels of community opportunity for women.

## Discussion

This analysis was the first to test an integrated gender-systems theory for the community-level influences on the hazard that a daughter experiences FGMC in a North African country that historically has adhered to a classic patriarchal family system (59). We examined to what extent norms opposing FGMC and more extra-familial opportunities for women in the community were associated with a girl’s lower risk of FGMC. We also assessed to what extent maternal opposition to FGMC was more strongly associated with the hazard of cutting a daughter in communities with more unfavorable FGMC norms and greater extra-familial opportunities for women. We leveraged rich data on FGMC from the 2014 Egypt DHS, which is well suited for multilevel analyses of community characteristics.

As expected, more unfavorable FGMC norms in the community were directly associated with a lower hazard that a daughter was cut (*H1*). However, extra-familial opportunities for women in the community were not directly associated with this hazard. Still, as expected, maternal opposition to FGMC was more negatively associated with the hazard that a daughter was cut in communities with more unfavorable FGMC norms and greater extra-familial opportunities for women (*H2*). These findings were robust to the inclusion of controls, and therefore, provide good support for a gender-systems theory of the multilevel influences on FGMC. Thus, mothers who oppose FGMC are less likely to have their daughter cut if they live in a community that more strongly opposes FGMC and offers more opportunity for women outside the family.

Results were mixed about there being a “tipping point” in community FGMC norms and women’s opportunities above which maternal opposition to FGMC reduces a daughter’s hazard of FGMC. In most age risk sets, the coefficient for the relationship of maternal opposition to FGMC with a daughter’s risk of FGMC was significant and negative, even in communities that strongly favored FGMC and had limited opportunities for women, relative to the average community in Egypt. Evidence supporting a “tipping point” was strongest only for the youngest age risk set (0–1), in which the probability of FGMC was very low.

Egypt provides a salient test case for our theory, being the country with the largest number of girls and women who have been cut and a persistently high prevalence of FGMC (11). In countries where more variation in the practice of FGMC exists, and thus a wider range of community normative contexts, we might expect an even stronger association of an unfavorable community gender system with daughter’s risk of FGMC. Yet, some distinctive features of the Egyptian context may make it difficult to generalize results across all countries where FGMC is practiced. Egypt has a strong national identity dominated by a single ethnic group and a single religion, although with visible minority groups (36). Thus, the nature of social identity and the relationship of FGMC to a sense of group belonging may differ in Egypt than in the multiethnic states that dominate sub-Saharan Africa. Moreover, although there is a large rural population in Egypt, the Egyptian economy is more diversified and less dependent on subsistence agriculture than are the economies of other countries where FGMC is common. The greater economic opportunity outside family-based agriculture, coupled with persistent elements of classic patriarchy that reinforce women’s economic dependence on male family members (59), may make expanded opportunities for women outside the family more consequential in Egypt than in other contexts (34). Comparative research to address these questions is warranted. Also, conditions surrounding the practice of FGMC in diaspora communities, where the maintenance of gender norms is tied to questions of cultural preservation and immigrant integration, may be distinct and require specific theorization.

### Limitations and implications for research

This analysis was not without limitations, which can inform recommendations for research. First, the cross-sectional design of the DHS limited our ability to establish the timing of community-level characteristics before the onset of a daughter’s risk of cutting. We addressed this limitation by constructing community-level factors using data from ever-married women 20–49 years, an older cohort than the one for which the FGMC outcome was observed. Still, if the composition of a community changed substantially or if families moved between a daughter’s birth and the survey, our community-level measures may have misrepresented the salient factors at the time of decision-making about FGMC. This measurement error would tend to reduce the precision of estimates and downwardly bias coefficients. Re-estimating final models without the oldest daughters (15–20 years) confirmed that the relationship of community FGMC norms with a daughter’s risk of FGMC was stronger. Thus, longitudinal surveys are needed to measure time-varying community characteristics before the onset of a daughter’s risk of FGMC.

Second, the DHS sample cluster was a proxy for “community.” The DHS is not designed for aggregation at the cluster level, and sample averages may produce imprecise estimates of true cluster characteristics, especially when the number of women per cluster is small and the intra-class correlation (ICCs) of the predictors is low (98). Depending on the characteristic being estimated, sample means also may produce biased estimates if the degree of precision is related to other community characteristics. In this sample, the mean cluster size was 24 ever-married women 20–49 years (range 1–127, SD 13). Estimated ICCs for community opportunities and community norms were .30 and .27, respectively, above the .20-level at which bias becomes unacceptable for the standard DHS cluster size of 25 respondents (98). Still, the average cluster size was small, so we cannot state definitively the level of the bias of cluster-level estimates without conducting simulations. Prior simulations have shown that the bias produced by using DHS sample clusters to estimate community characteristics is small (98). Surveys intended for multilevel analyses should draw separate within-community samples that are large enough to construct unbiased estimates of community-level characteristics that are measured by aggregating individual responses (99).

Third, a conceptual concern with using the DHS sample cluster as the measure of community is that the cluster may not capture the multiple social communities in which mother-daughter pairs are embedded (38). Network, spatial, and qualitative data to measure women’s overlapping activity spaces and social-group memberships would offer a more nuanced understanding of the relevant socio-contextual influences on a daughter’s risk of cutting.

Fourth, our measure for women’s community-level opportunities relied on indicators of schooling attainment, engagement in market work, and age at first residence with a spouse. These measures are important but may not capture the range of women’s local opportunities, nor women’s experience of opportunity. More contextualized community-level measures of women’s opportunities, or empowerment, may influence more strongly a daughter’s risk of FGMC.

Fifth, the community-level exposures of interest were correlated, such that communities tending to disfavor FGMC also tended to have more opportunities for women. Our theory of gender systems predicted some correlation among these measures and their distinct influences; still, distortion due to multi-collinearity may be a concern. In descriptive analyses, these community-level exposures had an estimated Pearson pairwise correlation of 0.64; however, in models using only one of the two community-level exposures, standard errors for the two community measures changed very little in these models, suggesting that collinearity was not a major problem. Thus, these constructs arguably were correlated but distinct components of a community gender system.

Finally, our analysis focused on two levels–the mother-daughter pair and the community. Yet, the influences of regional or national conditions warrant consideration. In sensitivity analyses, results for primary explanatory variables were robust to including region fixed effects in our final model. Since our primary interest was in community norms, community opportunity structures, and cross-level interactions, we retained the more parsimonious model without regional dummies. In future analyses, comparing individual, community, and national influences on a daughter’s FGMC risk would reflect a comprehensive ecological model of change.

### Implications for policies and programs

Our findings have several implications for policies and programs. Community-level opposition to FGMC had a direct association with daughter’s FGMC risk and strengthened the protective effect of maternal opposition to FGMC. This finding corroborates evidence from TOSTAN, a quasi-experimental intervention study of the relationship between a human-rights-based community empowerment and educational curriculum and girls’ risk of experiencing FGMC. In TOSTAN, communities were engaged in participatory dialogues and community-based activities grounded in the principles of human rights, democratic processes, and women’s health, among other topics. At the completion of these dialogues, communities deliberated and spearheaded collective decisions about abandoning harmful practices, including FGMC, and engaged neighboring communities to adopt the same new norms. The study showed evidence of diffusion of abandonment from program participants to non-participants and more unfavorable attitudes about FGMC and lower prevalence of cutting girls in intervention than comparison areas (100). Thus, interventions aimed at changing collective perceptions of FGMC—such as community pledges to reject FGMC—may have multiplicative impacts on daughters’ outcomes.

Community-level opportunities for women were not directly associated with daughter’s FGMC risk, but our results imply that improving opportunities for women may reduce the risk of FGMC among daughters whose mothers oppose the practice. Thus, future community-based interventions might consider joint activities that involve informal educational opportunities and training in income-generating activities for mothers. Future research should explore the joint influence of community FGMC norms among men on the risk of FGMC among daughters and whether engaging men in community-based programs would help to accelerate the abandonment of FGMC. Going forward, practitioners should consider intervention packages that address community gender norms specific to FGMC and that provide enabling resources to mothers, which empower them collectively to imagine alternatives to dependence on marriage in the life course trajectories of their daughters. Any effort to shift gender norms and opportunity structures will be most effective if it attempts to respect the values of local communities.

Despite the importance of community context, we also found that maternal opposition to FGMC was protective for daughters at most ages, even in communities that strongly opposed change. Thus, even where community-level interventions are infeasible, interventions focused on the beliefs of individuals may support change. Individual-level interventions might include provider support for non-cutting during clinic visits, or one-on-one support from outreach workers or religious leaders.

### Conclusion

Results provide good support for a gender-systems theory of the multilevel influences on FGMC. Integrated, multilevel interventions that address community-level gender norms about FGMC, community-level opportunities for women outside the family, and personal beliefs about FGMC among mothers of at-risk daughters may be needed for sustainable declines in the practice.

## Data Availability

This study is based on an analysis of secondary, deidentified data that is publicly available by request at the link, below.

https://dhsprogram.com/data/available-datasets.cfm

## Acknowledgements

Assistance from Kanghong Shao in data preparation is appreciated. This research was supported by research grant 1R21HD086762-01/02 (PIs Yount and Hayford). Funding also was received from the National Center on HIV/AIDS, Viral Hepatitis, STD, and TB Prevention, Centers for Disease Control and Prevention; Health Resources and Service Administration in support of the Health Policy Leadership Fellowship in the Satcher Health Leadership Institute at Morehouse School of Medicine; and Ohio State University’s Institute for Population Research (P2c-HD058484). This manuscript was prepared while Dr. Laurie James-Hawkins and Dr. Rose Grose were post-doctoral fellows in the Hubert Department of Global Health at Emory University.

